# The Genetic Architecture of Brain Structure and Function: A Data-Driven Interpretation Using genomICA

**DOI:** 10.1101/2025.04.07.25324950

**Authors:** Nicolò Trevisan, Lennart M. Oblong, Margo Raijmakers, Sourena Soheili-Nezhad, Yingjie Shi, Christian F. Beckmann, Emma Sprooten

## Abstract

Understanding the genetic basis of individual differences in human brain structure and function remains a major challenge. Traditional genome-wide association studies (GWAS) have identified numerous genetic variants associated with brain phenotypes, but their interpretation is complicated by polygenicity and pleiotropy. To address this, we applied genomICA, a data-driven multivariate method, to GWAS summary statistics of thousands of MRI-derived brain phenotypes from 33,224 UK Biobank participants. genomICA decomposes high-dimensional genetic data into independent components (ICs), capturing shared patterns of genetic influence across multiple traits. We identified 16 ICs, collectively explaining 39.2% of the variance. We describe each IC in detail here and in an online database (genomica.info). In addition to IC-brain and IC-genomic loadings, we explored the IC’s biological underpinnings through gene set enrichment and trait association analyses. Our results revealed that the generated components highlighted a diversity of neurobiological processes such as stress response (IC2), inflammation (IC5, IC15), glutamatergic signaling (IC8), lipid and semaphorin pathways (IC10), and circadian rhythms (IC12). In addition, some components reflect complex behavioral/lifestyle aspects such as diet and risk taking. genomICA offers a novel, data-driven framework for dissecting the complex genetic architecture of brain phenotypes, moving beyond univariate GWAS. This exploratory study provides a valuable resource for the imaging and genetic community, with all components and associated data available at genomICA.info, serving as a starting point for hypothesis generation and downstream analyses in brain health and disease.

## Introduction

The intricate biological mechanisms that underpin individual differences in brain structure and function remain a significant challenge in neuroscience. Magnetic resonance imaging (MRI) studies have consistently demonstrated that many brain phenotypes exhibit substantial heritability, indicating a strong genetic component to neural variability among individuals (Elliott et al., 2018; Eyler et al., 2011). Genome-wide association studies (GWAS) have been pivotal in identifying common genetic variants associated with a wide array of neuroimaging traits, and mental health conditions and behavioral phenotypes. However, the interpretation of GWAS results is complicated by the highly polygenic and pleiotropic nature of these traits, where numerous genetic variants contribute to multiple phenotypic traits with small effect sizes (Matoba et al., 2022; Wendt et al., 2020).

Traditional GWAS methodologies generate massive datasets of univariate statistics for millions of single nucleotide polymorphisms (SNPs), quantifying their individual associations with specific traits. While this approach has yielded valuable insights, testing each SNP-association separately falls short in capturing the complex mechanisms between genetic variants and the biological pathways they influence. The volume of GWAS data and the complex mechanisms they reflect, necessitates novel analytical frameworks that can distil these data into more interpretable components, potentially reflecting underlying biological processes.

Alternative methods for examining pleiotropic effects in GWAS data include genomic structural equation modeling (SEM) (Grotzinger et al., 2019), genomic Principal Component Analysis (PCA) (Fürtjes et al., 2023), and the Multivariate Omnibus Statistical Test (MOSTest) (Grotzinger et al., 2019). SEM requires the researcher to specify a model that describes how genetic variants influence multiple phenotypes, an assumption that may not always be justified and which can constrain the analysis. Although MOSTest combines SNP statistics across traits into a single component, it does not allow for disentangling multiple distinct patterns of pleiotropic effects. In contrast, genomic PCA is entirely data-driven; however, because its components are derived by maximizing the overall explained variance, they may mix distinct biological signals into a single factor. A data-driven approach that decomposes high-dimensional datasets into statistically independent components, therefore, offers a promising avenue to unravel the complex genetic architecture underlying brain phenotypes.

To address this challenge, we developed genomic Independent Component Analysis (genomICA). genomICA applies Independent Component Analysis (ICA) to GWAS summary statistics across a large number of phenotypes. This allows for the identification of independent patterns of genetic associations that are shared across multiple traits, effectively capturing pleiotropic effects into a smaller number of genome-wide factors or “independent genomic components”. We have previously shown the reproducibility of these genomic components when applied to MRI-derived brain phenotypes (Oblong et al., 2024). Our data-driven approach yields multiple independent genome-brain components and can handle thousands of phenotypes simultaneously, making it particularly suited for high-dimensional GWAS data.

Building upon this foundation, the present study applies genomICA again on thousands of brain MRI phenotypes from a large sample of 33,224 participants from the UK Biobank. To balance model complexity and capture a substantial proportion of the variance in the high-dimensional GWAS data, we focused on extracting 16 ICs. Here, we focus on describing and understanding the outcome of genomICA, i.e. towards a narrative of what these discovered independent genome-wide components may reflect at the level of genes, the molecular pathways, and brain morphology.

Our approach involves several key steps: First, we apply ICA to GWAS summary statistics of thousands of MRI-derived brain phenotypes to identify hidden genomic factors contributing to individual variability in brain phenotypes. Second, we investigate the neuroimaging phenotypes that exhibit substantial loadings on each IC to determine which brain MRI measures are most strongly represented by each component. Third, we investigate associations between these independent components and a comprehensive set of non-brain traits catalogued in the GWAS Atlas using Functional Mapping and Annotation (FUMA) (Watanabe et al., 2017). This allows us to explore how the identified genomic components relate to a broad spectrum of phenotypic outcomes, including behavioral and clinical traits. Fourth, we perform gene-level association analyses, followed by gene set enrichment analysis using Multi-marker Analysis of Genomic Annotation (MAGMA) (De Leeuw et al., 2015). This integrative approach enables us to connect genetic variation to biological pathways and processes, providing deeper insights into the genetic architecture of brain phenotypes.

In the following sections, we provide detailed exploratory narrative interpretations of each IC. For each IC, we discuss the significant genetic associations, the implicated genes and gene sets, the relevant biological pathways, the associated traits from the GWAS catalog, and the neuroimaging phenotypes that exhibit the strongest loadings. Our aim is to provide a comprehensive, data-driven characterization of each genome-brain component. This serves two primary purposes: to generate novel, data-driven, testable hypotheses about how specific patterns of genetic variation influence individual differences in brain structure and function, and to create a resource that allows researchers to select and utilize relevant components for downstream analyses, such as polygenic scoring in their own studies and datasets, including potential applications in stratification of clinical samples.

## Methods

### GWAS Data

We obtained GWAS summary statistics from the Oxford Brain Imaging Genetics (BIG40) database, which comprises results from over 4,000 GWAS analyses performed on MRI phenotypes within the UK Biobank (UKBB) cohort. ¹ For this study, we utilized the full sample of 33,224 UKBB participants to maximize statistical power and enhance the robustness of our findings. The MRI traits included metrics from T1-weighted MRI, diffusion MRI, susceptibility-weighted imaging (SWI), fluid-attenuated inversion recovery (FLAIR), task MRI, and quality control (QC) procedures. For detailed information on the GWAS pipeline and the specific MRI phenotypes analyzed, please refer to the original publications (Oblong et al., 2024) and the BIG40 database website (https://open.win.ox.ac.uk/ukbiobank/big40).

### Clumping

We applied genome-wide SNP clumping to reduce local SNP dependencies arising from linkage disequilibrium (LD) (Purcell et al., 2007). The smallest p-value of each SNP across the GWASs of 2240 brain traits was selected for clumping. A genomic window size of 250 kilobases, a lead variant p-value threshold of p < 0.001, and an LD-threshold of r² < 0.5 were used as clumping parameters. LD was estimated using the European reference panel from the 1000 Genomes project (Durbin et al., 2010). This procedure reduced the total number of genome-wide SNPs from n = 17,103,079 to 1,032,967. This approach reduces the impact of LD on local SNP-to-SNP correlations while retaining brain-related lead variants within each LD block.

### genomICA

We applied genomICA to GWAS summary statistics of MRI phenotypes as in (Oblong et al., 2024). By concatenating the standardised GWAS regression beta’s—representing genome-wide SNP effect sizes—across all brain measures, we constructed a brain-wide genome-wide matrix with dimensions m × n, where m = 2,240 MRI traits and n = 1,029,040 clumped SNPs. This matrix captures the effect sizes of genetic variants on brain phenotypes.

Using the Multivariate Exploratory Linear Optimised Decomposition into Independent Components (MELODIC) algorithm (Beckmann & Smith, 2004), we decomposed this high-dimensional data into ICs, each representing a latent genomic component influencing brain structure and function. Each IC comprises a vector of SNP loadings and a corresponding vector of MRI loadings. This structure allows us to explore and tentatively interpret the latent genomic components represented by each component.

We disabled the global signal removal and variance normalization options in MELODIC to preserve the biological significance of the allelic effect magnitudes. Detailed methodological parameters and computational considerations are described in our previous publications, to which we refer interested readers for further information.

Using genomICA, we extracted 16 ICs, which cumulatively explained 39.2% of the total variance in the GWAS summary statistics. This proportion of explained variance is consistent with our previous methodological work which demonstrated robust and reproducible results when capturing this amount of variance (by 10 components using more strictly clumped data). This choice thus allowed us to replicate a comparable level of explained variance to our prior work, even when analyzing a larger set of SNPs in this study due to less stringent clumping.

### FUMA

We went on to investigate all 16 ICs genetic associations with traits catalogued in the GWAS Catalog and identified relevant genes and gene sets using Functional Mapping and Annotation (FUMA) of GWAS data. FUMA is a web-based platform that facilitates the annotation, prioritization, and mapping of genetic variants to genes and biological pathways, enhancing the interpretability of GWAS results (Watanabe et al., 2017).

#### SNP Selection and Genomic Loci Identification

For each IC, we identified significant SNPs by transforming their (normalised) loadings (z-scores) into P-values for further analysis. To define independent significant SNPs representing genomic loci, we utilized the FUMA platform with a sample size of 33,224. SNPs were considered genome-wide significant if they had P-values below 5 × 10⁻⁸. The reference panel population was based on the 1000 Genomes Phase 3 European (EUR) dataset. Genomic risk loci were established by merging lead SNPs and their associated LD blocks when they were within 250 kilobases (kb) of each other. These parameters are consistent with FUMA’s default settings for locus definition.

#### Functional Annotation and Gene Mapping

Candidate SNPs were determined by including all SNPs in LD with the independent significant SNPs at r² ≥ 0.6 threshold, including those not directly genotyped but present in the reference panel. FUMA mapped these SNPs to genes using Positional Mapping by assigning SNPs to genes based on their physical location within or proximal to a gene with a 20kb window.

#### Trait Association Analysis

To explore phenotypic associations, we cross-referenced the genomic risk loci with the GWAS Catalog to identify traits that were previously associated with these loci in the literature. To interpret the results we excluded brain-MRI traits from the output (since they were a given and they overcrowded the output), as well as traits associated with oncological diseases.

By integrating the results from FUMA, we aimed to elucidate the significance of the ICs derived from genomICA, linking genetic variation to functional pathways and other complex traits. This approach provides a comprehensive framework for describing and interpreting the genetic architecture underlying brain structure and function.

### MAGMA

#### Gene-Level Association Analysis with MAGMA

To further characterize the genetic architecture underlying the independent components (ICs), we conducted gene-level association analyses using Multi-marker Analysis of Genomic Annotation (MAGMA) (De Leeuw et al., 2015). Unlike our previous locus-based approaches, this method incorporated all SNPs associated with the ICs, thereby providing a more comprehensive view of gene-level associations. We executed MAGMA from the command line rather than via the FUMA web interface, which allowed for greater transparency and flexibility in parameter selection and results interpretation. For gene mapping, we applied MAGMA’s ‘snp-wise=top’ model with an annotation window extending 35 kilobases upstream and 10 kilobases downstream of each gene; SNPs falling within these boundaries were assigned to the corresponding gene, thereby capturing potential regulatory regions that may influence gene expression. We opted for the ‘snp-wise=top’ option because it does not rely on assumptions linking absolute p-values to local LD structure, a particularly important consideration given that our output may not conform to the assumptions underlying standard GWAS summary statistics.

#### Gene Set Enrichment Analysis

To identify biological pathways and functional categories associated with the genes linked to our ICs, we conducted gene set enrichment analysis using MAGMA. This approach assesses, for each gene set, whether the genes within the set show more significant associations than genes that are not in the gene set, taking into account gene size and LD-structure. We utilized the Human Molecular Signatures Database (MSigDB) collections, specifically the “C5: Ontology Gene Sets,” which include 16,107 gene sets derived from Gene Ontology (GO) terms encompassing biological processes, cellular components, and molecular functions.

By integrating gene-level associations with curated gene sets, we aimed to uncover overrepresented biological pathways and processes that may underlie the genomic signals captured by the ICs. This analysis provides insights into the putative biological relevance of the genomic factors influencing brain structure and function.

## Results

Our analyses revealed that these components encapsulate a diverse array of biological processes and mechanisms. They highlight roles in extracellular matrix organization, myelination, immune system function, neural development, lipid metabolism, circadian rhythm regulation. The neuroimaging phenotypes associated with each IC tend to correspond to distinct contributions of MRI modalities, brain regions and tissues, including cortical and subcortical areas, white matter tracts, and regions implicated in motor control and cognitive functions.

All results, including detailed visualizations and data for each component, are also available at genomICA.info, serving as a resource for further exploration by the scientific community.

### Component 1 (IC1): Polygenic Architecture of Brain Structure and Lifestyle

IC1 emerges as a broadly polygenic factor without pronounced enrichment in any pre-curated gene sets, suggesting wide-ranging genetic influences on brain structure. This component also appears influenced by lifestyle factors, highlighting traits such as smoking and alcohol consumption. The most significant SNP of IC1 is located in a locus mapping to VCAN (Rahmani et al., 2006), which encodes an extracellular matrix component important for cell adhesion. CDC42, a Rho GTPase, is identified through locus mapping to genes within this component. Located on chromosome 1, CDC42 plays a pivotal role in orchestrating the actin cytoskeleton and is integral to neuronal morphogenesis, axonal guidance, and synapse formation (Etienne-Manneville & Hall, 2002). These functions are underpinned by its ability to regulate cell polarity and membrane dynamics in developing neurons. In the same locus on chromosome 1 we also found WNT4, identified by both locus gene mapping and via MAGMA gene analysis, which further supports a role in structural and organizational processes within the nervous system. WNT4, as part of the Wnt signaling pathway, mediates cell fate specification, cell-to-cell adhesion, and tissue patterning, all of which are vital to neurodevelopment and neural circuit maintenance (Khan & Verma, 2025). ACTR1B, located on chromosome 2, is identified as the most significant gene in the MAGMA analysis for this component, being also present in the locus mapping to gene. ACTR1B encodes subunits of dynactin, which is crucial for retrograde axonal transport and interacts with the actin cytoskeleton (Schroer, 2004). This further reinforces the component’s link to cytoskeletal dynamics and neuronal structure.

The MRI brain measures that load most heavily on IC1 predominantly reflect diffusion metrics in white matter tracts, indicative of underlying differences in microstructural properties. These data, when integrated with the genetic findings, point to a possible collective influence of actin-mediated cytoskeletal remodeling and cell adhesion mechanisms on white matter organization. This component appears highly distributed across the genome, consistent with a polygenic architecture and non-specific mechanisms such as cell adhesion.

### Component 2 (IC2): Myelination and Stress Response

IC2 is a much less widespread polygenic component, showing several clear peak loci. IC2 tentatively reflects myelin-associated pathways, stress-response mechanisms, and neurodegenerative processes. The Manhattan plot for this component shows a notable peak on chromosome 17, mapping to, CRHR1 (corticotropin-releasing hormone receptor 1) on chromosome 17 linking IC2 to stress regulation (Perrelli et al., 2024). As a mediator of the hypothalamic-pituitary-adrenal (HPA) axis, CRHR1 modulates systemic stress responses. The same locus also involves MAPT (microtubule-associated protein tau), which is related to neurodegeneration. This is also underscored by the FUMA trait analysis, showing that this locus has been previously associated with multiple neurological diseases such as Parkonsin’s and Alzheimer’s disease.

**Figure 1.**
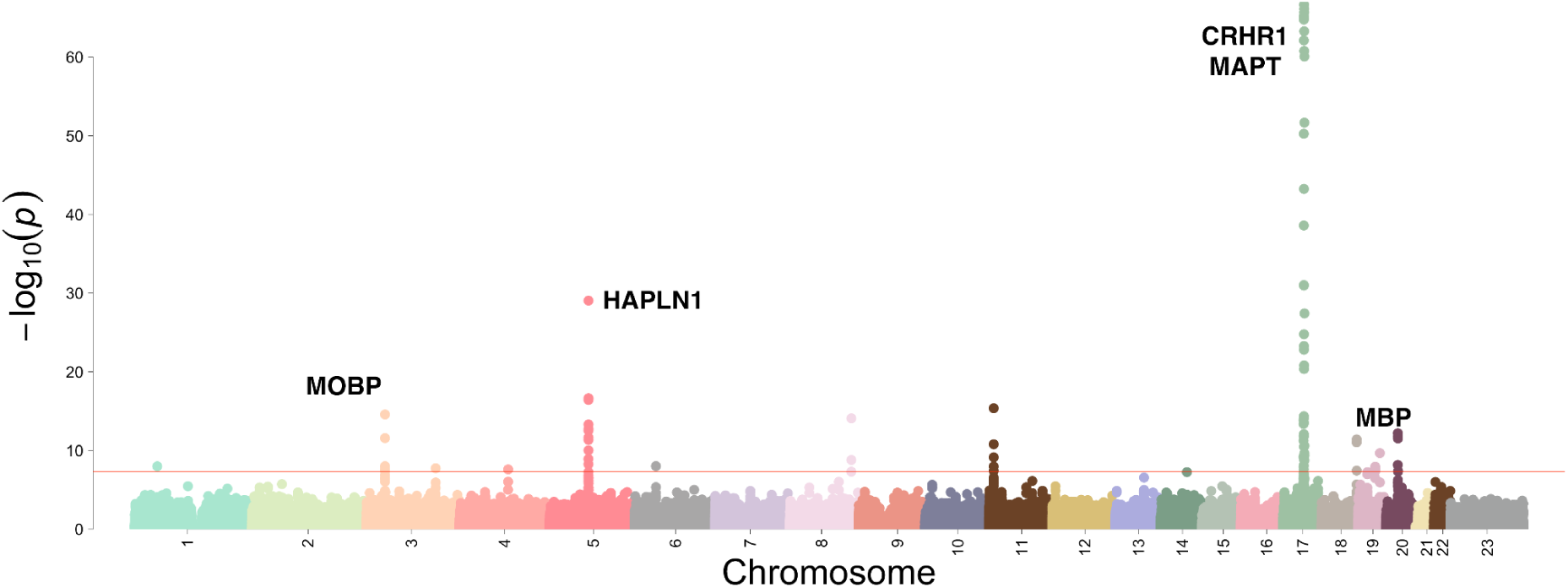
Manhattan plot for IC2. Each point represents a SNP, with the x-axis showing chromosomal position (1–23) and the y-axis showing the −log10(*p*) values derived from the genomICA loadings. The red horizontal line marks the genome-wide significance threshold (5×10⁻⁸). The most notable peak occurs on chromosome 17, with additional minor signals on chromosomes 3, 5 and 18. All component-specific plots and full data are available at genomICA.info, where researchers can further explore these and other ICs in detail.

The second most significant locus maps to HAPLN1 (hyaluronan and proteoglycan link protein 1) on chromosome 5, which encodes a protein critical for extracellular matrix (ECM) stability. By maintaining ECM integrity, HAPLN1 supports oligodendrocyte function and myelin structure, aligning with the component’s focus on myelination (Oohashi et al., 2015).

Two other IC2 loci each map to myelin proteins; MOBP (myelin-associated oligodendrocyte basic protein) and MBP (myelin basic protein) on chromosomes 3 and 18, respectively. MOBP stabilizes the myelin membrane, while MBP ensures myelin compaction, both vital for efficient axonal conduction (Aggarwal et al., 2013).

**Figure 2.**
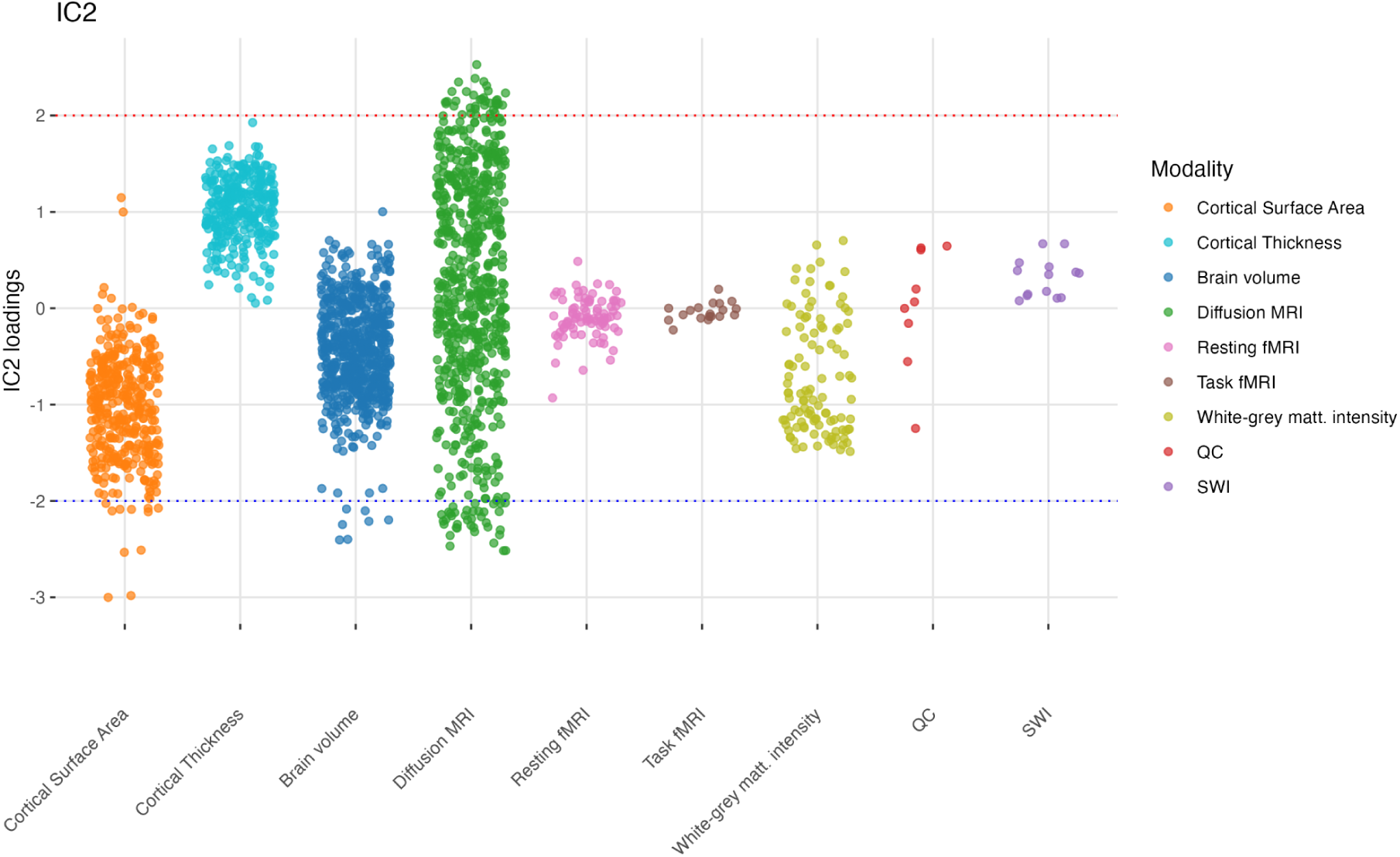
Distribution of MRI trait loadings for Independent Component 2 (IC2) across different imaging modalities. This scatter plot displays the loadings of IC2 on individual MRI-derived traits, categorized by modality. Each point represents a single MRI trait, with its position on the y-axis indicating the magnitude and direction of its loading on IC2. The x-axis groups the traits by MRI modality. The plot illustrates that Diffusion MRI traits exhibit the highest positive loadings on IC2, while Cortical Surface Area traits show negative loadings, suggesting a differential relationship of IC2 with these distinct MRI modalities.

MRI traits prominently loading on IC2 include total brain volume, total surface area, ventral diencephalon and striatum volumes, and multiple diffusion-based measures. While diffusion metrics reflect myelinated fibre pathways, other traits such as brain volume may capture broader structural or neurodevelopmental correlates. The ventral diencephalon is interesting as a top region for this locus, as it contains the hypothalamus, thus part of the HPA-axis.

**Figure 3.**
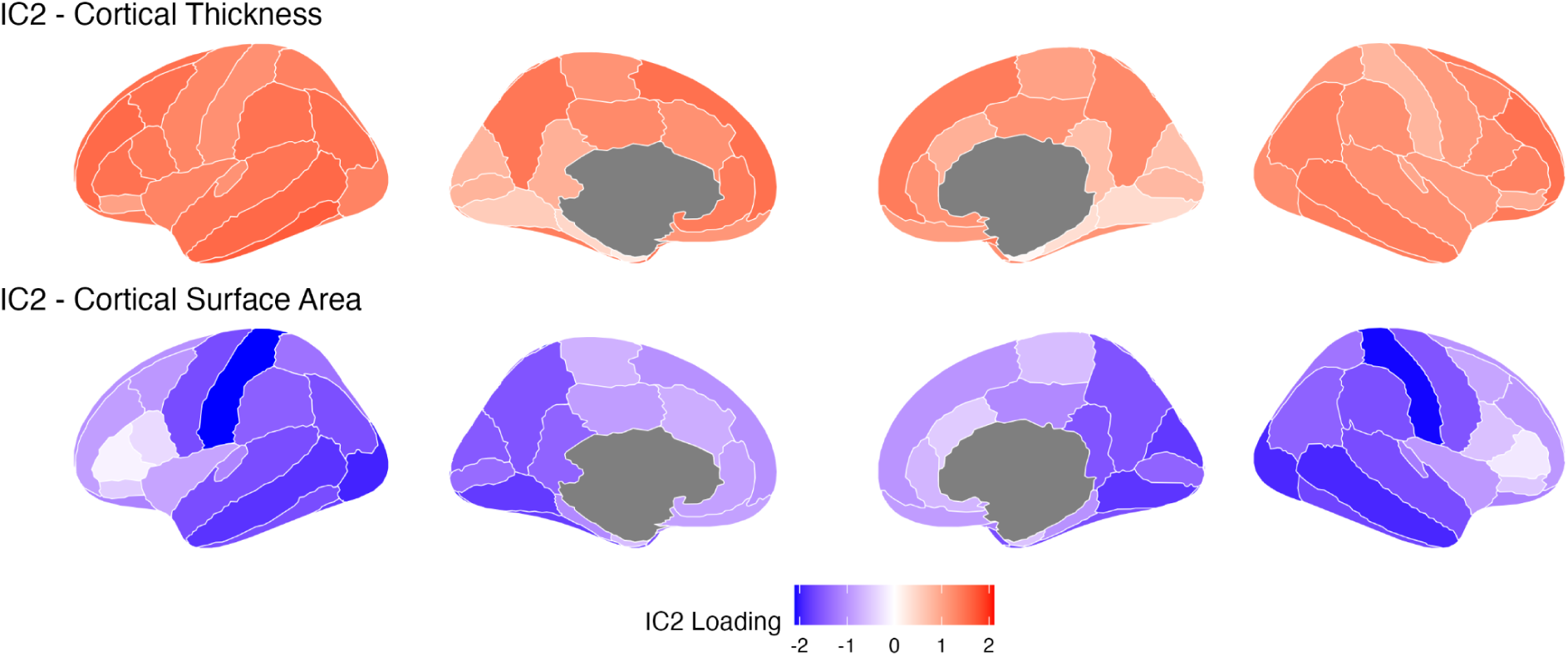
Cortical thickness and cortical surface area loadings for IC2 (DKT atlas). Regions in red/orange indicate higher positive loadings on IC2. The intensity scale corresponds to the magnitude of the IC loading for this specific component and MRI trait.

Finally, the GWAS catalog analyses resulted in hundreds of non-MRI traits previously associated with aforementioned loci, including neurodegeneration, psychiatric conditions, lifestyle factors, and blood biomarkers suggest IC2 integrates myelination, stress pathways, and environmental influences, shaping brain structure and integrity.

### Component 3 (IC3): Cortical Development

IC3 is distinguished by a strong genetic signal evidenced by a huge peak on chromosome 5. This locus also emerged in IC1, but is much more prominent for IC3. The locus maps to two genes involved in extracellular matrix (ECM) organization and cell adhesion: VCAN (versican) and HAPLN1 (hyaluronan and proteoglycan link protein 1). Both genes are also identified as the most strongly significant genes in both locus mapping to genes and MAGMA statistics. VCAN is a large chondroitin sulfate proteoglycan critical for cellular adhesion, migration, and tissue remodeling, particularly during neural development. HAPLN1 contributes to ECM stability by cross-linking hyaluronan with proteoglycans, facilitating structural integrity in the developing brain.

The associated gene set GOBP_NK_T_CELL_DIFFERENTIATION suggests that immune-modulatory processes, including the development and activation of NK and T cells, may intersect with brain development in this component. From a phenotypic standpoint, HP_APLASIA_HYPOPLASIA_OF_THE_NASAL_SEPTUM indicates developmental anomalies potentially sharing underlying pathways with brain development.

MRI traits loading onto IC3 show contrasting effects across cortical volume and thickness versus subcortical volumes. Taken together, IC3 appears to represent core genetic underpinnings of cortical development, with strong evidence from a prominent peak on chromosome 5 driven by VCAN and HAPLN1.

### Component 4 (IC4): Polygenic Regulation of MAPK Signaling and Neuronal Plasticity

IC4 is characterized by a relatively sparse genetic profile, indicating a highly distributed polygenic architecture. Yet its associated gene sets point toward phosphatase-mediated regulation of the mitogen-activated protein kinase (MAPK) pathway. In particular, GOMF_MAP_KINASE_TYROSINE_SERINE_THREONINE_PHOSPHATASE_ACTI VITY and GOMF_PROTEIN_TYROSINE_THREONINE_PHOSPHATASE_ACTIVITY highlight the dephosphorylation of MAPK components, which is instrumental in modulating cell growth, differentiation, and survival (Shaul & Seger, 2007). Within the nervous system, MAPK activity underpins numerous processes central to neuronal plasticity, including learning, memory formation, and synaptic remodeling. The high-level involvement of phosphatases, such as dual-specificity phosphatases (DUSPs), underscores the fine-tuned control of phosphorylation states necessary for precise neural function (Thomas & Huganir, 2004).

From an MRI traits perspective, IC4 reflects volumetric, thickness, and diffusion-weighted imaging measures, though specific trait correlations are lacking. The absence of robust single-gene drivers or well-defined trait associations points to a widespread, polygenic influence on neural microstructure or functional connectivity. Consequently, IC4 is likely reflective of broad, systemic modulation of brain imaging phenotypes through fundamental cell signaling processes, implying that variations in phosphatase-driven MAPK regulation can have diffuse but meaningful effects on brain structure and function.

### Component 5 (IC5): Inflammation and Neurotrophic Support

IC5 is notable for a prominent genetic locus on chromosome 6 overlapping the human leukocyte antigen (HLA) region, as clearly seen by a spike in the Manhattan plot and identified both via locus gene mapping and MAGMA gene analysis. This dominant signal from the HLA region strongly underscores the component’s immunological underpinnings. The HLA complex encodes molecules pivotal for antigen presentation, facilitating immune surveillance and contributing to inflammatory processes that can influence neural tissue integrity. Beyond this immunological signature, BDNF (brain-derived neurotrophic factor) on chromosome 11 emerges as a key contributor, though its signal is less prominent than the HLA region (Lima Giacobbo et al., 2019). BDNF is essential for neuronal growth, survival, and synaptic plasticity, and has been repeatedly implicated in learning and memory formation, particularly within medial temporal lobe structures. There are also indications that BDNF is protective against neuroinflammation (Porter & O’Connor, 2022).

Additional genes contributing to IC5 include GMNC (geminin coiled-coil domain containing) on chromosome 3, a chromatin-binding factor that governs cell cycle progression and DNA replication, and SCFD2 (Sec1 family domain containing) on chromosome 4 (both of these genes are identified via locus gene mapping and MAGMA analysis), which participates in vesicular exocytosis and, consequently, neurotransmitter release and synaptic communication. The recurrent involvement of VCAN and HAPLN1 on chromosome 5, also observed in other components, further emphasizes the role of extracellular matrix organization in immune and trophic signals.

MRI traits loading on IC5 are predominantly localized to the temporal lobes—particularly diffusion and T1-weighted intensity measures. Given that the temporal lobe is rich in BDNF expression and highly susceptible to inflammatory processes, these converging signals may point to an integrated mechanism by which immune-related pathways and neurotrophic factors shape regional brain structure and function. Altogether, IC5 may thus encapsulate an interplay between immunogenetic factors (notably HLA loci), neurotrophic support (via BDNF), and extracellular matrix remodeling (via VCAN and HAPLN1), culminating in modulations of temporal lobe microstructure and function. The genetic architecture of IC5 is largely driven by the strong signal from the HLA region on chromosome 6.

### Component 6 (IC6): Genetic Associations with Motor Pathways and Unclear Functional Interpretation

IC6 features a prominent genetic locus on chromosome 1, encompassing genes such as MR1 (major histocompatibility complex, class I–related) and STX6 (syntaxin 6). MR1 is involved in presenting microbial metabolites to immune cells, while STX6 plays a role in intracellular transport. Another locus on chromosome 6 includes CENPW (centromere protein W), important for chromosome segregation during cell division. These genes were identified through locus mapping (Kwon et al., 2007).

MRI traits with high loadings on IC6 are primarily related to motor pathways, including the corticospinal tract, precentral gyrus, brainstem, and cerebellum.

Interestingly, gene set enrichment analysis highlighted GOBP_HINDLIMB_MORPHOGENESIS as a significant association for IC6. However, the connection between this developmental gene set and the other genetic and neuroimaging findings for IC6 is not immediately apparent, and the functional meaning of this component remains uncertain.

In conclusion, IC6 captures a combination of genetic associations and motor-related MRI traits. While we have identified suggestive genes and a gene set, a definitive interpretation of the underlying biological processes represented by IC6 is currently lacking.

### Component 7 (IC7): Lifestyle and Ventricles

IC7 is characterized by a prominent genetic signal on chromosome 16, with FBXO31 (F-box protein 31) emerging as the most significant gene in both MAGMA and locus mapping analyses. FBXO31, may have a role in regulating the cell cycle as well as dendrite growth and neuronal migration (Vadhvani et al., 2013). A secondary locus on chromosome 7 implicates GNA12 (G protein subunit alpha 12), a regulator of Gα12/13 signaling pathways. GNA12 is essential for neuronal migration and synaptic organization during development. Its oncogenic roles are well-documented, but its intersection with neurodevelopmental pathways highlights potential pleiotropy in IC7’s architecture (Moers et al., 2008).

Trait associations for IC7 include ventricular volume, tau pathology, and many lifestyle factors (e.g., sleep, BMI, physical activity, smoking, diet). MRI measures with strongest loadings localize to the ventricles, thalamus, and fornix, regions sensitive to cerebrospinal fluid (CSF) dynamics and structural atrophy. Ventricular enlargement, a nonspecific biomarker of brain atrophy, correlates with aging and neurodegeneration. Lifestyle factors may indirectly influence these phenotypes through metabolic or inflammatory pathways, though genetic causality remains unproven.

### Component 8 (IC8): Glutamatergic Signaling and Cortical Development

IC8 is driven by genes linked to zinc and calcium homeostasis, intersecting with glutamatergic synaptic function and cortical development. All genes are significant in both locus gene mapping and MAGMA analyses. SLC39A8 (solute carrier family 39 member 8, chromosome 4), a zinc transporter critical for systemic and neuronal zinc regulation, indirectly modulates synaptic plasticity and NMDA receptor activity (Lichten & Cousins, 2009). Variants in SLC39A8 are associated with neurodevelopmental risks (Poulter et al., 2013). MSRB3 (methionine sulfoxide reductase B3, chromosome 12), a zinc-dependent enzyme, mitigates oxidative damage to proteins, preserving mitochondrial integrity and neuronal viability. DOC2A (double C2-like domain-containing alpha, chromosome 16), a calcium sensor mostly expressed in brain tissues, regulates neurotransmitter release in glutamatergic synapses (Courtney et al., 2018).

Notably, chromosome 16 contributes PPP4C (protein phosphatase 4 catalytic subunit) and YPEL3 (yippee-like 3), both located in the 16p11.2 region. PPP4C has an important role for cortical development, while YPEL3 modulates cellular senescence and growth arrest, processes tied to developmental stability. The 16p11.2 locus is a known hotspot for structural polymorphisms linked to autism and intellectual disability, aligning with IC8’s neurodevelopmental focus (Toyo-oka et al., 2008).

**Figure 4.**
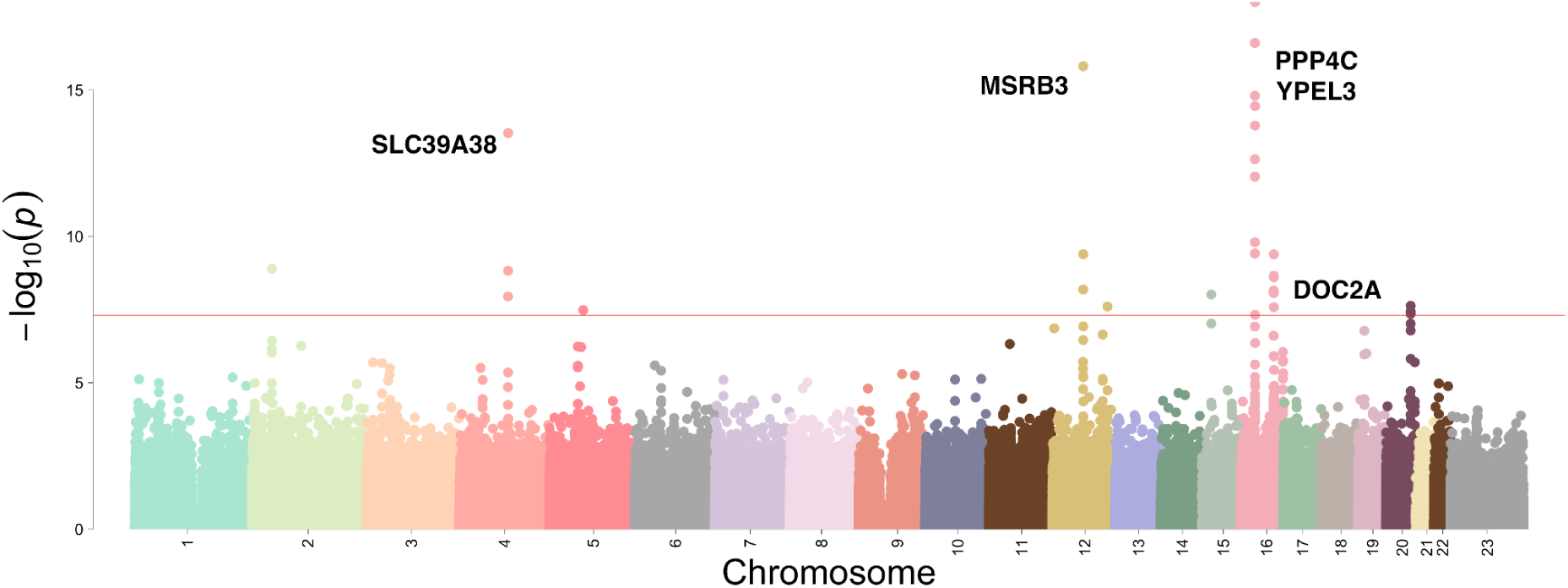
Manhattan plot for IC8. Each point represents a SNP, with the x-axis showing chromosomal position (1–23) and the y-axis showing the −log10(*p*) values derived from the genomICA loadings. The red horizontal line marks the genome-wide significance threshold (5×10⁻⁸). Notable peaks occur on chromosome 16, with additional signals on chromosomes 4 and 12, indicating a polygenic component.

**Figure 5.**
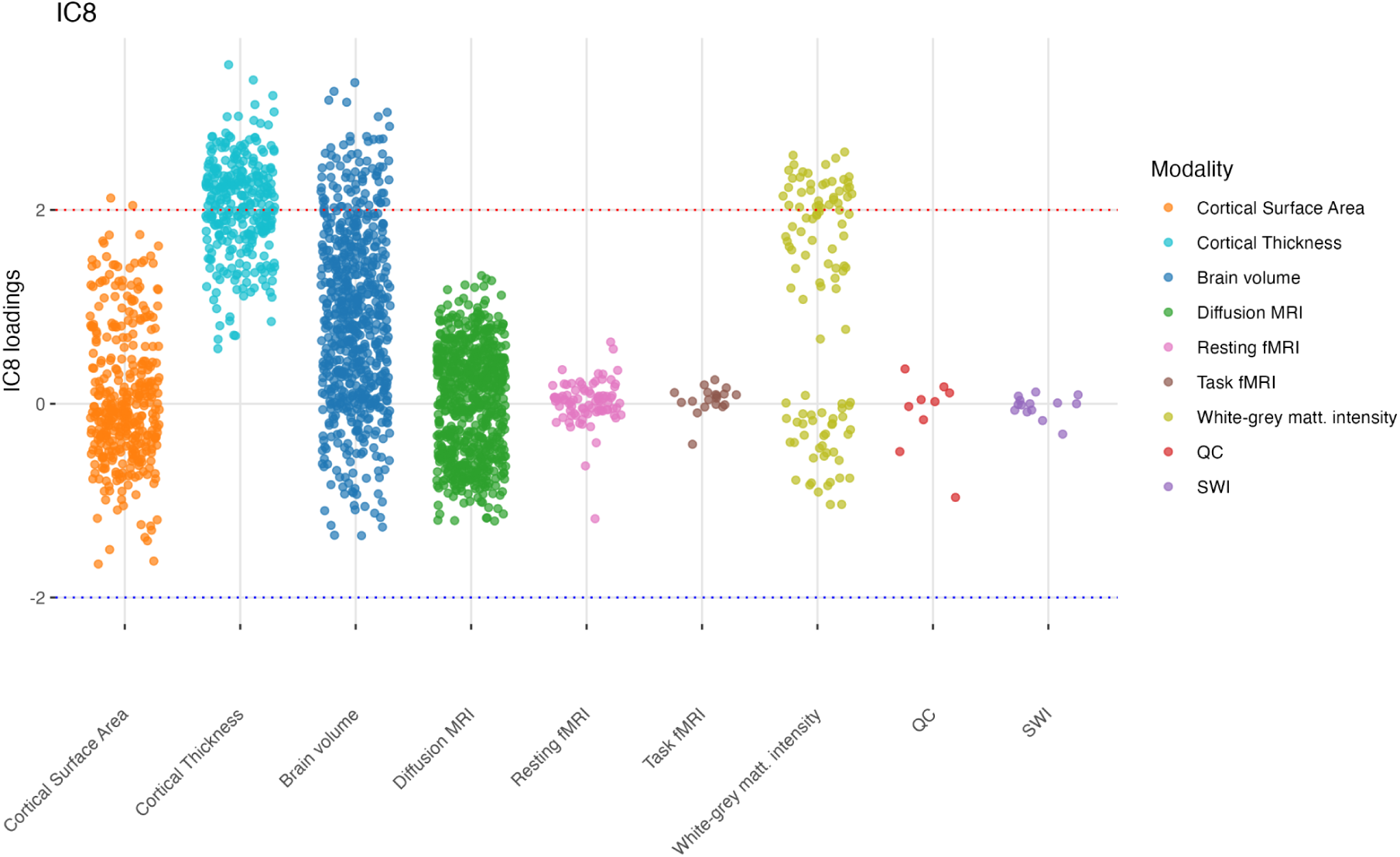
Distribution of MRI trait loadings for Independent Component 8 (IC8) across different imaging modalities. This scatter plot displays the loadings of IC8 on individual MRI-derived traits, categorized by modality. Each point represents a single MRI trait, with its position on the y-axis indicating the magnitude and direction of its loading on IC2. The x-axis groups the traits by MRI modality. The plot reveals that Cortical Thickness traits exhibit the highest positive loadings on IC8, while Cortical Surface Area traits tend to show negative loadings. Brain Volume and White-grey matter intensity contrast traits also display positive loadings, suggesting a modality-specific pattern of association between IC8 and brain structure.

MRI traits loading onto IC8 predominantly highlight cortical thickness variations in glutamatergic-rich regions, such as the prefrontal and sensory cortices. Trait associations include lifestyle factors (diet, alcohol intake, smoking) and ventricular volume. While lifestyle behaviors may influence zinc/calcium availability, their genetic interplay with IC8 loci remains observational. Ventricular enlargement, a marker of brain atrophy, may reflect broader disruptions in ion homeostasis or developmental pathways.

**Figure 6.**
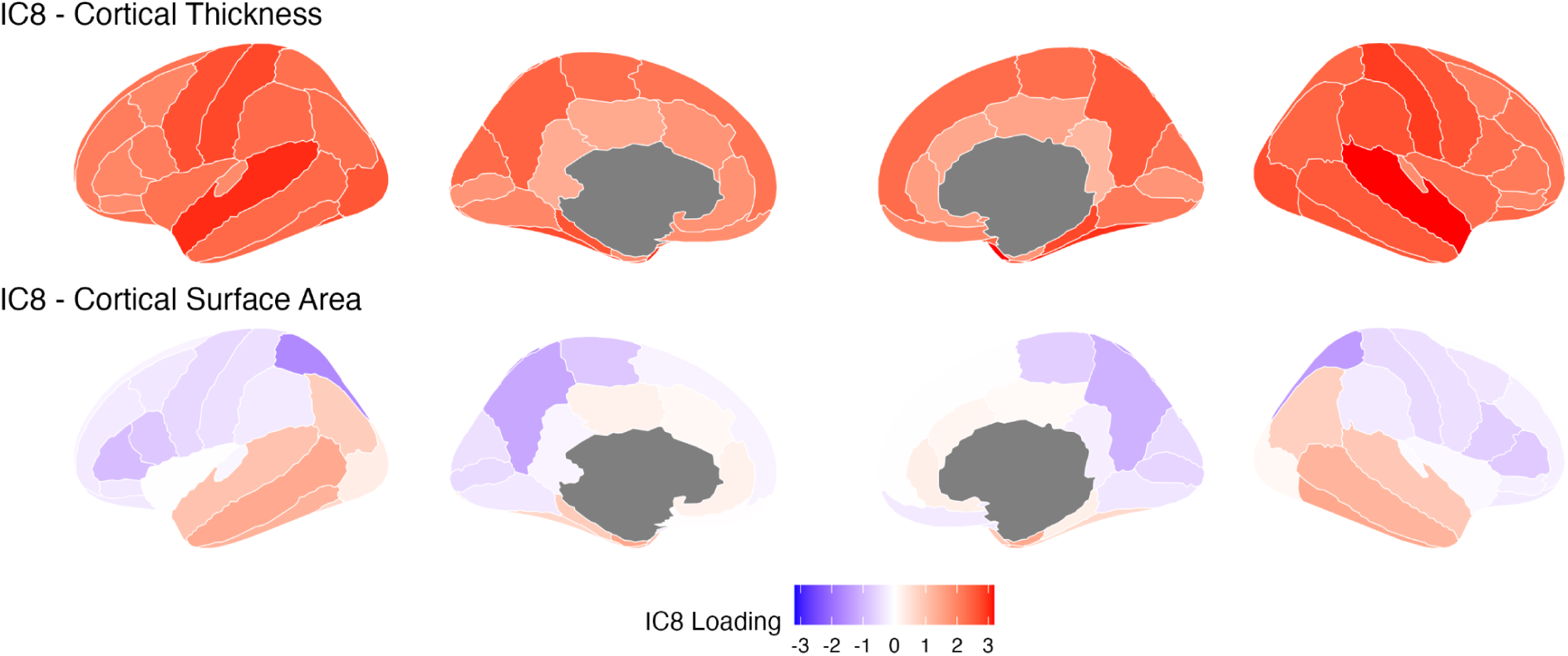
Cortical thickness (a) and cortical surface area (b) loadings for IC8 (DKT atlas). Regions in red/orange indicate higher positive loadings on IC8. The intensity scale corresponds to the magnitude of the IC loading for this specific component and MRI trait.

IC8’s genetic architecture integrates zinc/calcium signaling (SLC39A8, DOC2A), redox balance (MSRB3), and developmental regulation (PPP4C, YPEL3), underscoring a convergence of synaptic, metabolic, and structural mechanisms in neurodevelopment and cortical integrity.

### Component 9 (IC9): Genetic Control of Brain Asymmetry and Lateralized Cognitive Functions

IC9 is marked by a significant locus on chromosome 14 encompassing DAAM1 (dishevelled-associated activator of morphogenesis 1), which showed the most significant gene-level association in MAGMA analysis. DAAM1 is integral to Wnt/β-catenin signaling, a core pathway governing cell fate, tissue patterning, and cytoskeletal dynamics, and drives actin cytoskeleton rearrangements required for cell polarity and axonal guidance (Habas et al., 2001).

Also significant are ZIC1 and ZIC4 (zinc-finger proteins of the cerebellum 1 and 4) on chromosome 3. These genes regulate transcriptional programs for midline patterning and cerebellar development, and are implicated in structural asymmetries (Aruga, 2004).

Trait associations for IC9 highlight mathematics and language aptitudes, as well as systemic left–right body asymmetries. MRI traits show differential loadings in occipital regions versus language-associated cortices, and higher loadings for right hemisphere thickness and intensity.

In sum, IC9 captures a genetic architecture governing brain lateralization, strongly driven by DAAM1 and supported by ZIC1 and ZIC4, all genes playing roles in tissue patterning and axon guidance, and aligning with lateralized brain functions and structural features.

### Component 10 (IC10): Lipid and Semaphorin Pathways in Myelination

IC10 reflects a polygenic profile, with loci converging on lipid metabolism and axon guidance pathways. Three major genes define its lipid-related dimension: GAL3ST1 (chromosome 22), ABHD12 (chromosome 20), and SLC27A3 (chromosome 1), showing significance in both locus gene mapping and MAGMA statistics.

GAL3ST1 catalyzes sulfation of galactosylceramide, generating sulfatides critical for myelin stability and neuronal conductivity. ABHD12 hydrolyzes lysophosphatidylserine (lyso-PS), a pro-inflammatory lipid implicated in synaptic pruning and microglial regulation; mutations here are linked to PHARC syndrome (polyneuropathy, hearing loss, ataxia) (El-Fasakhany et al., 2001). SLC27A3 mediates long-chain fatty acid transport, influencing neuronal and glial metabolic homeostasis (Kazantzis & Stahl, 2012).

The GOCC_SEMAPHORIN_RECEPTOR_COMPLEX gene-set underscores semaphorin signaling, a pathway guiding axonal pathfinding and neural migration. Semaphorins (e.g., SEMA3A) and receptors (e.g., neuropilins, plexins) regulate cytoskeletal dynamics, shaping neural circuitry in development and adulthood (Takahashi et al., 1999).

Trait associations span blood lipid markers (LDL, HDL), insulin resistance, dietary patterns, and mental health indices, suggesting lipid-semaphorin crosstalk may influence metabolic and cognitive outcomes. MRI traits highlight gray–white matter intensity contrast in the nucleus accumbens and basal ganglia, regions where myelin integrity depends on sulfatide synthesis (GAL3ST1). Diffusion metrics in the cerebellum and brainstem align with semaphorin-mediated axon guidance defects.

In summary, IC10 captures genetic interplay between lipid metabolism (ABHD12, GAL3ST1, SLC27A3) and semaphorin signaling, potentially affecting myelination, neural connectivity, and systemic metabolism. While polygenic, lipid-related loci dominate its architecture.

### Component 11 (IC11): Polygenic Basis of Risk-Taking Behavior and Associated Traits

IC11 represents a highly polygenic component, characterized by a broad spectrum of trait associations and a notable, though not singular, concentration of genetic signal on chromosome 8. Due to its polygenic nature, pinpointing specific causal genes is challenging, and the listed genes from MAGMA analysis should be considered as potential candidates within a much larger polygenic landscape. Despite the lack of clear single-gene drivers, the consistent association of five independent loci with risk-taking behavior is particularly interesting. This suggests that IC11, through its complex polygenic architecture, may influence fundamental biological processes that have downstream consequences on behavioral traits related to impulsivity, reward processing, and decision-making.

Multiple loci are also associated with blood pressure and educational attainment. While the co-occurrence of such traits alongside risk-taking might appear disparate, it is plausible that shared, broadly acting biological mechanisms contribute to these diverse phenotypes. For example, systemic processes like stress response dysregulation or chronic low-grade inflammation could influence both metabolic health and neural circuits involved in risk assessment and impulsive actions. It is important to note that attributing specific roles to individual genes within this polygenic component is speculative at this stage. Genes like FDFT1, DEFB cluster, MSRA, and SOX7 on chromosome 8 are mentioned as examples of genes within the region, but their individual contributions to risk-taking or other IC11 traits remain to be determined and are likely to be small within the overall polygenic context. Similarly, the presence of developmental genes like ZIC1 and ZIC4 on chromosome 3 highlights the potential for developmental origins of these pleiotropic effects, but again, within a complex polygenic framework.

In summary, IC11 is best characterized as a highly polygenic component associated with a wide array of traits, with a noteworthy association with risk-taking behaviors. This component likely reflects a complex and distributed genetic architecture impacting diverse biological systems, manifesting in variations in brain volumes and cortical thickness, and influencing both physiological and behavioral traits. While the chromosome 8 peak is observed, the polygenic nature of IC11 makes it difficult to attribute causality to specific genes. Further research is needed to disentangle the complex polygenic architecture of IC11 and to understand the broader biological mechanisms that link its diverse trait associations, with a particular focus on elucidating the polygenic underpinnings of its influence on risk-taking behavior. This interpretation remains tentative and emphasizes the need for caution in over-interpreting the role of individual candidate genes within this highly polygenic component.

### Component 12 (IC12): Genetic Regulation of Circadian Rhythms and Sleep

IC12 is dominated by a huge genetic peak on chromosome 15, indicating a major genetic influence on this component from this region. Our automated gene mapping pipeline did not pinpoint specific genes within this locus. However, further manual investigation of the chromosome 15 peak reveals that it is located close to GPR176, a gene known to be a key regulator of the circadian clock. This suggests that GPR176, or potentially other genes in this locus, are likely major contributors to the chromosome 15 signal and to IC12 overall (Doi et al., 2016).

In addition to this chromosome 15 signal found through manual investigation, our automated analyses identified associations with DDN (dendrin, chromosome 12) and VAX1 (ventral anterior homeobox 1, chromosome 10), both significant in locus gene mapping and MAGMA analyses. DDN is important for synaptic plasticity during sleep, and VAX1 is crucial for the development of the suprachiasmatic nucleus (SCN) (Hoffmann et al., 2021; Neuner-Jehle et al., 1996). MAGMA analysis also identified LINC02915 (chromosome 15), a long non-coding RNA located within the dominant chromosome 15 locus, though its precise role in circadian rhythms is not yet well-established.

MRI traits loading on IC12 include total brain volume and cortical surface area, with a specific pattern in sensorimotor cortices. Trait associations include various sleep-related phenotypes like insomnia and sleep disturbance.

In summary, IC12 reflects a strong genetic influence from the chromosome 15 locus (suggestively linked to GPR176 through manual investigation), alongside contributions from genes identified by our automated pipeline that are involved in synaptic plasticity and SCN development. This component captures a genetic architecture shaping both brain structure and sleep-wake regulation, with a major signal originating from the chromosome 15 locus and pointing towards circadian rhythm mechanisms.

### Component 13 (IC13): Polygenic Convergence on Myelination, Cytoskeletal Organization, and Lipid Homeostasis

IC13 reflects a polygenic architecture with loci converging on pathways critical for myelination, cytoskeletal organization, and lipid homeostasis. Locus mapping identified genes such as GAL3ST1 (chromosome 22), DAAM1 (chromosome 14), MOBP (chromosome 3), and SEC14L4 (chromosome 22).

GAL3ST1 is involved in sulfatide biosynthesis, essential for myelin stability. MOBP is a structural component of myelin. DAAM1 regulates the cytoskeleton. SEC14L4 is hypothesized to be involved in lipid transport (El-Fasakhany et al., 2001).

Trait associations with IC13 include neuroticism, intelligence, and circulating sulfatide levels. MRI traits load most strongly on white matter structures.

Despite these suggestive locus-based gene associations, IC13’s polygenic nature and the lack of strong single gene drivers make a definitive interpretation challenging. The observed connections to myelination, cytoskeleton, and lipid pathways, and the trait associations, remain tentative and require further investigation to understand the underlying biological mechanisms.

### Component 14 (IC14): Immune-Metabolic Interactions in Cortical Development

IC14 is characterized by a lack of strong individual gene or locus associations. Instead, its most informative features come from gene set enrichment analysis, which reveals significant associations with pathways related to immune regulation, lipid metabolism, and developmental signaling. While MAGMA analysis prioritizes genes like HLA-DRA, ARHGEF12, NUP210L, and TPM3, the signal for IC14 appears to be more broadly distributed across gene sets than concentrated in specific loci.

The most significant gene sets for IC14 include GOBP_NOTCH_SIGNALING_PATHWAY, GOBP_NEGATIVE_REGULATION_OF_FATTY_ACID, and GOBP_LEUKOTRIENE_D4_BIOSYNTHETIC_PROCESS. These suggest a convergence on Notch signaling, crucial for cortical layering, and metabolic-immune crosstalk involving fatty acid metabolism and inflammatory leukotrienes (Lasky & Wu, 2005; Nian & Hou, 2022).

Consistent with these biological themes, MRI traits loading on IC14 show volumetric differences in frontal and temporal cortices. Interestingly, cortical thickness measures exhibit a clear left-right hemispheric split in their loadings on IC14, suggesting potential lateralized effects.

In conclusion, IC14 is primarily a gene set driven component, highlighting the importance of immune-metabolic and developmental pathways, particularly Notch signaling, fatty acid metabolism, and leukotriene biosynthesis, in shaping cortical structure. The observed left-right asymmetry in cortical thickness adds another layer of complexity, suggesting potential lateralized roles for these pathways in cortical development. Further investigation should focus on the interplay of these biological processes in cortical organization.

### Component 15 (IC15): Inflammation and Lipid Metabolism Influencing White Matter

IC15 highlights a genetic network influencing white matter organization via intersecting immune and lipid metabolic pathways. Among its notable features is a significant locus within the major histocompatibility complex (MHC) on chromosome 6, encompassing all of the HLA genes integral to immune response modulation. This HLA region signal is evident in both locus gene mapping and MAGMA statistics. Beyond this immunogenetic core, ENPP6 (ectonucleotide pyrophosphatase/phosphodiesterase 6) on chromosome 4, also significant in both analyses, plays a pivotal role in myelin homeostasis through sphingolipid metabolism, thereby supporting efficient signal conduction in the central nervous system (Morita et al., 2016). The adjacent involvement of DEFB135 and DEFB136 (defensin beta 135 and 136) on chromosome 8, emphasizes the significance of innate immune effectors in mitigating pathogens and maintaining neuroimmune balance (Xu & Lu, 2020).

Additionally, LPAR1 (lysophosphatidic acid receptor 1) on chromosome 9, significant in both analyses, involved in lipid signaling, myelination and immune regulation. LPAR1, a G-protein–coupled receptor, modulates cell proliferation, migration, and inflammatory responses via lysophosphatidic acid. The link to FTH1 (ferritin heavy chain 1) on chromosome 11, also significant in both analyses, further integrates iron homeostasis with neural integrity and oxidative stress defense (Ward et al., 2014).

In line with these molecular indicators, MRI traits reveal strong loadings on intracellular volume fraction from diffusion-weighted MRI, suggesting that these loci collectively affect white matter cellular composition, possibly reflecting alterations in oligodendrocyte density or myelin structure.

Trait associations—including dietary preferences (e.g., fish and pork consumption), cholesterol metrics, white blood cell counts, and inflammatory markers—underscore how variations in immune and metabolic pathways can shape both systemic health and white matter microstructure.

Altogether, the genetic signal for IC15 is driven by a prominent locus in the HLA region on chromosome 6 and supported by contributions from other loci involved in lipid metabolism and immune function. IC15 captures a multifaceted genetic framework wherein immune response, lipid metabolism, and iron regulation converge to influence white matter integrity, offering insight into potential susceptibility factors for neuroinflammatory and demyelinating conditions.

### Component 16 (IC16): Genetic Regulation of Cell Differentiation

Component 16 (IC16): Genetic Regulation of Cell Differentiation - Centriole Assembly Focus

IC16 is characterized by a very dominant genetic locus on chromosome 15, a signal shared with IC12, underscoring recurrent genetic factors in neurodevelopmental processes. This chromosome 15 locus includes LINC02915, a long intergenic non-protein coding RNA, which may play a regulatory role in neurodevelopment (Mercer et al., 2008).

Gene-set analysis reveals a strong association with GOBP_DE_NOVO_CENTRIOLE_ASSEMBLY, implicating centriole and cilia function in neurogenesis and neural circuit maturation. Centrioles are microtubule-organizing centers essential for cell division and ciliogenesis, processes critical for neural progenitor proliferation and migration in the developing central nervous system (Breslow & Holland, 2019).

MRI traits show heterogeneous loadings across regions involved in motor control, cognitive function, and metabolic regulation. Trait associations include progressive supranuclear palsy, Creutzfeldt-Jakob disease, and metabolic/immune indices.

In summary, IC16 captures a genetic influence that is chiefly driven by the chromosome 15 locus and its involvement in de novo centriole assembly, underscoring a key biological pathway likely critical for neural development.

## Discussion

This exploratory study employed genomICA, a powerful multivariate method, to dissect the complex genetic architecture of brain structure and function using GWAS summary statistics from thousands of neuroimaging-derived phenotypes in the UK Biobank. Our data-driven approach decomposed this high-dimensional genetic data into 16 ICs, each representing a constellation of genetic influences on a range of brain phenotypes. These components, collectively explaining a substantial portion of the variance in the GWAS data, offer a novel framework for understanding the pleiotropic and polygenic nature of brain traits.

The identified ICs revealed a rich tapestry of biological processes and pathways shaping brain variability. Several components converged on fundamental neurobiological themes, providing compelling insights into the genetic underpinnings of brain organization. For instance, IC2, IC13, and IC15 consistently implicated myelination, highlighting the critical role of myelin integrity in shaping brain structure and function (Fields, 2008). These components pointed to specific genes involved in sulfatide biosynthesis (GAL3ST1), myelin structure (MOBP), and lipid metabolism (ABHD12, SEC14L4), underscoring the complex interplay of genetic factors required for efficient myelination (Aggarwal et al., 2013; El-Fasakhany et al., 2001; Kazantzis & Stahl, 2012). Furthermore, the association of these components with diffusion MRI metrics in white matter tracts and broader brain volumes reinforces the functional relevance of these genetic pathways to macroscale brain organization.

Inflammation and immune processes emerged as another prominent theme, particularly in IC5, IC6, IC14, and IC15. The distinct signal from the HLA region in IC5 and IC15, coupled with the involvement of genes like MR1 and STX6 in IC6, strongly suggests that immunogenetic variation plays a significant role in shaping brain phenotypes (Ransohoff & Engelhardt, 2012). These components also implicated genes involved in neuroinflammation (e.g., LPAR1, HLA-DRA) and innate immunity (DEFB135/136), suggesting a complex interplay between systemic immune responses and brain microstructure (Xu & Lu, 2020). The association of IC5 with temporal lobe phenotypes, a region known for its susceptibility to inflammatory processes and rich in BDNF expression (Tokuyama et al., 2000), further strengthens this interpretation. BDNF, beyond its neurotrophic roles, has also been implicated in modulating neuroinflammatory responses (Porter & O’Connor, 2022; Shirayama et al., 2002), suggesting a potential protective or regulatory role in the context of IC5.

Several components, including IC3, IC7, IC8 and IC9, highlighted the importance of neurodevelopmental processes. These components implicated genes involved in cortical development (VCAN, HAPLN1, GFAP, PPP4C), cell differentiation (KLF3, NFIX), cytoskeletal dynamics (DAAM1, ACTR1B), and synaptic function (DDN, DOC2A). The enrichment of gene sets related to ECM organization, cell adhesion, and centriole assembly further underscored the developmental origins of brain structural variability (Habas et al., 2001; Schroer, 2004). Notably, IC9 specifically captured genetic influences on brain lateralization, implicating genes like DACT1 and ZIC4, and aligning with trait associations related to language and mathematical abilities, as well as structural features like falx cerebri calcification (Corballis, 2017).

Interestingly, lifestyle factors appeared to be linked to the influence of several components, particularly IC1, IC10, and IC11. Traits such as smoking, alcohol consumption, diet, and BMI were associated with these components, suggesting that environmental and behavioral factors are associated with the underlying genetic architecture that shape brain phenotypes. While the directionality of these associations requires further investigation, it highlights the potential for gene-environment correlations in influencing brain structure and function (Sprooten et al., 2022). IC11, despite its high polygenicity and lack of strong single gene signals, was notably associated with risk-taking behaviors, suggesting a complex polygenic basis for behavioral traits related to impulsivity and decision-making.

The application of genomICA offers several methodological advantages over traditional univariate GWAS approaches. By leveraging the inherent pleiotropy present in the GWAS summary statistics of multifactorial brain traits, where each SNP’s effect is assessed across many brain phenotypes, genomICA effectively distills the vast GWAS data of thousands of traits into more interpretable, independent components. This data-driven approach avoids the need for pre-defined models or assumptions about the relationships between genetic variants and phenotypes, allowing for the discovery of novel and unexpected patterns of genetic influence. Furthermore, genomICA’s ability to handle thousands of phenotypes simultaneously makes it particularly well-suited for analyzing high-dimensional GWAS summary statistics, providing a comprehensive view of the genome-brain architecture (Fürtjes et al., 2023; Grotzinger et al., 2019).

Despite these strengths, several limitations should be acknowledged. Our study relies on GWAS summary statistics, which inherently limits our ability to infer causality and necessitates caution in interpreting the directionality of observed associations. The interpretation of highly polygenic components, such as IC1, IC4, and IC11, remains challenging, as pinpointing specific causal genes within a broad genetic landscape is difficult. While gene set enrichment analysis provides valuable insights into biological pathways, these analyses are based on existing annotations and may not capture the full complexity of gene function or all relevant molecular processes. Furthermore, the explained variance of 39.2% by 16 components, while consistent with previous work, indicates that a substantial portion of the genetic variance in brain phenotypes remains unexplained, suggesting the involvement of additional genetic factorsthat are not yet reliably captured by the available GWAS. Larger discivery sample sizes will likely allow expanding on the numbers of ICs that can be derived reliably (Oblong et al., 2024). The 16 independent components we identified likely represent a subset of the total number of independent genetic mechanisms influencing brain structure and function. Our previous work suggests that with the current sample size, extracting 16 components captured a reliable portion of the multivariate variance. However, it is plausible that each of these components may still represent a combination of multiple underlying biological mechanisms, and that further independent components remain to be discovered within the substantial portion of variance (approximately 60%) not explained by our 16-component model. Adding to these limitations, we acknowledge that while our interpretations are guided by objective, data-driven tools, the narrative descriptions of each component ultimately involve a degree of subjectivity and are susceptible to reverse inference. Therefore, while we have provided tentative narrative interpretations for each component, further functional validation is crucial to confirm their biological relevance and refine our understanding of the underlying mechanisms.

Looking ahead, it is important to view this study as an initial, exploratory step in deciphering the complex genetic architecture of brain structure and function using genomICA. The identified ICs and their associated genetic loci, genes, and pathways offer a rich source of testable hypotheses about the genetic architecture of brain structure and function. Similar to the early applications of Independent Component Analysis in neuroimaging, where the functional significance of independent components was initially interpreted in a data-driven and somewhat tentative manner (Biswal & Ulmer, 1999; Damoiseaux et al., 2006), our interpretations of the genomICA components are intended to be hypothesis-generating. Researchers can utilize these components for downstream analyses, such as polygenic risk scoring in independent datasets, including clinical samples, to investigate the role of these genomic factors in brain disorders and behavioral phenotypes. Just as the names and interpretations of fMRI-derived networks have evolved and become more refined as neuroscientific knowledge has accumulated, we anticipate that the narratives and biological understanding of these genome-brain components will similarly evolve with future research and validation. The availability of all results at genomICA.info further enhances the accessibility and utility of this resource for the scientific community.

Although we already established replication of the ICs in independent samples (Oblong et al., 2024), future research should focus on further validating these findings in independent cohorts and employing functional genomics approaches to elucidate the causal mechanisms underlying the identified genome-brain components. A key avenue for maximizing the impact of these genomICA components is to make them publicly available as a valuable resource for the scientific community. By providing open access to these components, we aim to empower researchers to readily utilize them for a wide range of downstream analyses in their own datasets, including the development of polygenic component scores (PCS) to predict brain phenotypes and related clinical outcomes. To further enrich the understanding and utility of these components, we also warmly invite researchers to explore the narratives and associated data available at genomICA.info and to contribute their diverse expertise and insights to complement and refine our initial interpretations. We believe that such collaborative, community-driven efforts will be invaluable in fully unlocking the biological significance of these genome-brain components and maximizing their impact on future neuroscience research. Integrating genomICA with other omics data, such as epigenomics and transcriptomics, could provide a more comprehensive understanding of the biological pathways mediating the genetic effects on brain phenotypes. Furthermore, exploring the clinical relevance of these components, particularly in the context of psychiatric and neurological disorders, holds significant promise for advancing precision medicine approaches in neuroscience.

In conclusion, this study demonstrates the power of genomICA to help to dissect the complex genetic architecture of brain structure and function. We have provided a novel, data-driven framework and starting point for further understanding the pleiotropic and polygenic nature of brain phenotypes. Our approach offers a valuable resource for future research and potentially paving the way for a more in depth understanding of the genetic basis of brain health and disease.

## Data and code availability

Complete data visualization is available at https://genomica.info/. The results of the present study were derived from the following resources available in the public domain: https://open.win.ox.ac.uk/ukbiobank/big40/. The code used to generate the genomic components generated by this study are found at https://github.com/LennartOblong/GenomicICA. The genomic components will be available at https://genomica.info/.

## Author contributions

N.T.: Conceptualization, Methodology, Data curation, Data analysis, Writing (Original Draft, review and editing, Visualization. L.O.: Conceptualization, Methodology, Data curation, Writing (Review and editing). M.R.: Data curation, Data analysis, Writing (Review and editing). S.S.: Writing (Review and editing). Y.S.: Writing (Review and editing). C.B.: Writing (Review and editing). E.S.: Conceptualization, Methodology, Data analysis, Writing (Review and editing).

## Data Availability

All data produced in the present study are available upon request to the authors
and also available online at https://genomica.info/
The code is available at https://github.com/LennartOblong/GenomicICA
The results of the present study were derived from the following resources available
in the public domain: https://open.win.ox.ac.uk/ukbiobank/big40/.

https://github.com/LennartOblong/GenomicICA.

https://open.win.ox.ac.uk/ukbiobank/big40/

## References

Aggarwal, S., Snaidero, N., Pähler, G., Frey, S., Sánchez, P., Zweckstetter, M., Janshoff, A., Schneider, A., Weil, M.-T., Schaap, I. A. T., Görlich, D., & Simons, M. (2013). Myelin membrane assembly is driven by a phase transition of myelin basic proteins into a cohesive protein meshwork. PLoS Biology, 11(6), e1001577. 10.1371/journal.pbio.1001577

Aruga, J. (2004). The role of *Zic* genes in neural development. Molecular and Cellular Neuroscience, 26(2), 205–221. 10.1016/j.mcn.2004.01.004

Beckmann, C. F., & Smith, S. M. (2004). Probabilistic Independent Component Analysis for Functional Magnetic Resonance Imaging. IEEE Transactions on Medical Imaging, 23(2), 137–152. 10.1109/TMI.2003.822821

Biswal, B. B., & Ulmer, J. L. (1999). Blind Source Separation of Multiple Signal Sources of fMRI Data Sets Using Independent Component Analysis. Journal of Computer Assisted Tomography, 23(2), 265.

Breslow, D. K., & Holland, A. J. (2019). Mechanism and regulation of centriole and cilium biogenesis. Annual Review of Biochemistry, 88, 691–724. 10.1146/annurev-biochem-013118-111153

Corballis, M. C. (2017). The Evolution of Lateralized Brain Circuits. Frontiers in Psychology, 8. 10.3389/fpsyg.2017.01021

Courtney, N. A., Briguglio, J. S., Bradberry, M. M., Greer, C., & Chapman, E. R. (2018). Excitatory and inhibitory neurons utilize different Ca2+ sensors and sources to regulate spontaneous release. Neuron, 98(5), 977. 10.1016/j.neuron.2018.04.022

Damoiseaux, J. S., Rombouts, S. A. R. B., Barkhof, F., Scheltens, P., Stam, C. J., Smith, S. M., & Beckmann, C. F. (2006). Consistent resting-state networks across healthy subjects. Proceedings of the National Academy of Sciences, 103(37), 13848–13853. 10.1073/pnas.0601417103

De Leeuw, C. A., Mooij, J. M., Heskes, T., & Posthuma, D. (2015). MAGMA: Generalized Gene-Set Analysis of GWAS Data. PLOS Computational Biology, 11(4), e1004219. 10.1371/journal.pcbi.1004219

Doi, M., Murai, I., Kunisue, S., Setsu, G., Uchio, N., Tanaka, R., Kobayashi, S., Shimatani, H., Hayashi, H., Chao, H.-W., Nakagawa, Y., Takahashi, Y., Hotta, Y., Yasunaga, J., Matsuoka, M., Hastings, M. H., Kiyonari, H., & Okamura, H. (2016). Gpr176 is a Gz-linked orphan G-protein-coupled receptor that sets the pace of circadian behaviour. Nature Communications, 7(1), 10583. 10.1038/ncomms10583

Durbin, R. M., Altshuler, D., Durbin, R. M., Abecasis, G. R., Bentley, D. R., Chakravarti, A., Clark, A. G., Collins, F. S., De La Vega, F. M., Donnelly, P., Egholm, M., Flicek, P., Gabriel, S. B., Gibbs, R. A., Knoppers, B. M., Lander, E. S., Lehrach, H., Mardis, E. R., McVean, G. A., … The Translational Genomics Research Institute. (2010). A map of human genome variation from population-scale sequencing. Nature, 467(7319), 1061–1073. 10.1038/nature09534

El-Fasakhany, F. M., Uchimura, K., Kannagi, R., & Muramatsu, T. (2001). A Novel Human Gal-3-*O*-Sulfotransferase. Journal of Biological Chemistry, 276(29), 26988–26994. 10.1074/jbc.M100348200

Elliott, L. T., Sharp, K., Alfaro-Almagro, F., Shi, S., Miller, K. L., Douaud, G., Marchini, J., & Smith, S. M. (2018). Genome-wide association studies of brain imaging phenotypes in UK Biobank. Nature, 562(7726), 210–216. 10.1038/s41586-018-0571-7

Etienne-Manneville, S., & Hall, A. (2002). Rho GTPases in cell biology. Nature, 420(6916), 629–635. 10.1038/nature01148

Eyler, L. T., Prom-Wormley, E., Panizzon, M. S., Kaup, A. R., Fennema-Notestine, C., Neale, M. C., Jernigan, T. L., Fischl, B., Franz, C. E., Lyons, M. J., Grant, M., Stevens, A., Pacheco, J., Perry, M. E., Schmitt, J. E., Seidman, L. J., Thermenos, H. W., Tsuang, M. T., Chen, C.-H., … Kremen, W. S. (2011). Genetic and Environmental Contributions to Regional Cortical Surface Area in Humans: A Magnetic Resonance Imaging Twin Study. Cerebral Cortex, 21(10), 2313–2321. 10.1093/cercor/bhr013

Fields, R. D. (2008). White matter in learning, cognition and psychiatric disorders. Trends in Neurosciences, 31(7), 361–370. 10.1016/j.tins.2008.04.001

Fürtjes, A. E., Arathimos, R., Coleman, J. R. I., Cole, J. H., Cox, S. R., Deary, I. J., De La Fuente, J., Madole, J. W., Tucker-Drob, E. M., & Ritchie, S. J. (2023). General dimensions of human brain morphometry inferred from genome-wide association data. Human Brain Mapping, 44(8), 3311–3323. 10.1002/hbm.26283

Grotzinger, A. D., Rhemtulla, M., De Vlaming, R., Ritchie, S. J., Mallard, T. T., Hill, W. D., Ip, H. F., Marioni, R. E., McIntosh, A. M., Deary, I. J., Koellinger, P. D., Harden, K. P., Nivard, M. G., & Tucker-Drob, E. M. (2019). Genomic structural equation modelling provides insights into the multivariate genetic architecture of complex traits. Nature Human Behaviour, 3(5), 513–525. 10.1038/s41562-019-0566-x

Habas, R., Kato, Y., & He, X. (2001). Wnt/Frizzled activation of Rho regulates vertebrate gastrulation and requires a novel Formin homology protein Daam1. Cell, 107(7), 843–854. 10.1016/s0092-8674(01)00614-6

Hoffmann, H. M., Meadows, J. D., Breuer, J. A., Yaw, A. M., Nguyen, D., Tonsfeldt, K. J., Chin, A. Y., Devries, B. M., Trang, C., Oosterhouse, H. J., Lee, J. S., Doser, J. W., Gorman, M. R., Welsh, D. K., & Mellon, P. L. (2021). The transcription factors SIX3 and VAX1 are required for suprachiasmatic nucleus circadian output and fertility in female mice. Journal of Neuroscience Research, 99(10), 2625–2645. 10.1002/jnr.24864

Kazantzis, M., & Stahl, A. (2012). Fatty Acid transport Proteins, implications in physiology and disease. Biochimica et Biophysica Acta, 1821(5), 852–857. 10.1016/j.bbalip.2011.09.010

Khan, M., & Verma, L. (2025). Crosstalk between signaling pathways (Rho/ROCK, TGF-β and Wnt/β-Catenin Pathways/PI3K-AKT-mTOR) in Cataract: A Mechanistic Exploration and therapeutic strategy. Gene, 149338. 10.1016/j.gene.2025.149338

Kwon, M.-S., Hori, T., Okada, M., & Fukagawa, T. (2007). CENP-C Is Involved in Chromosome Segregation, Mitotic Checkpoint Function, and Kinetochore Assembly. Molecular Biology of the Cell, 18(6), 2155–2168. 10.1091/mbc.E07-01-0045

Lasky, J. L., & Wu, H. (2005). Notch Signaling, Brain Development, and Human Disease. Pediatric Research, 57(7), 104–109. 10.1203/01.PDR.0000159632.70510.3D

Lichten, L. A., & Cousins, R. J. (2009). Mammalian zinc transporters: Nutritional and physiologic regulation. Annual Review of Nutrition, 29, 153–176. 10.1146/annurev-nutr-033009-083312

Lima Giacobbo, B., Doorduin, J., Klein, H. C., Dierckx, R. A. J. O., Bromberg, E., & de Vries, E. F. J. (2019). Brain-Derived Neurotrophic Factor in Brain Disorders: Focus on Neuroinflammation. Molecular Neurobiology, 56(5), 3295–3312. 10.1007/s12035-018-1283-6

Matoba, N., Love, M. I., & Stein, J. L. (2022). Evaluating brain structure traits as endophenotypes using polygenicity and discoverability. Human Brain Mapping, 43(1), 329–340. 10.1002/hbm.25257

Mercer, T. R., Dinger, M. E., Sunkin, S. M., Mehler, M. F., & Mattick, J. S. (2008). Specific expression of long noncoding RNAs in the mouse brain. Proceedings of the National Academy of Sciences of the United States of America, 105(2), 716–721. 10.1073/pnas.0706729105

Moers, A., Nürnberg, A., Goebbels, S., Wettschureck, N., & Offermanns, S. (2008). Gα12/Gα13 Deficiency Causes Localized Overmigration of Neurons in the Developing Cerebral and Cerebellar Cortices. Molecular and Cellular Biology, 28(5), 1480–1488. 10.1128/MCB.00651-07

Morita, J., Kano, K., Kato, K., Takita, H., Sakagami, H., Yamamoto, Y., Mihara, E., Ueda, H., Sato, T., Tokuyama, H., Arai, H., Asou, H., Takagi, J., Ishitani, R., Nishimasu, H., Nureki, O., & Aoki, J. (2016). Structure and biological function of ENPP6, a choline-specific glycerophosphodiester-phosphodiesterase. Scientific Reports, 6(1), 20995. 10.1038/srep20995

Neuner-Jehle, M., Denizot, J.-P., Borbély, A. a., & Mallet, J. (1996). Characterization and sleep deprivation-induced expression modulation of dendrin, a novel dendritic protein in rat brain neurons. Journal of Neuroscience Research, 46(2), 138–151. 10.1002/(SICI)1097-4547(19961015)46:2<138::AID-JNR2>3.0.CO;2-I

Nian, F.-S., & Hou, P.-S. (2022). Evolving Roles of Notch Signaling in Cortical Development. Frontiers in Neuroscience, 16. 10.3389/fnins.2022.844410

Oblong, L. M., Soheili-Nezhad, S., Trevisan, N., Shi, Y., Beckmann, C. F., & Sprooten, E. (2024). Principal and independent genomic components of brain structure and function. *Genes*, Brain and Behavior, 23(1), e12876. 10.1111/gbb.12876

Oohashi, T., Edamatsu, M., Bekku, Y., & Carulli, D. (2015). The hyaluronan and proteoglycan link proteins: Organizers of the brain extracellular matrix and key molecules for neuronal function and plasticity. Experimental Neurology, 274, 134–144. 10.1016/j.expneurol.2015.09.010

Perrelli, M., Goparaju, P., Postolache, T. T., Del Bosque-Plata, L., & Gragnoli, C. (2024). Stress and the CRH System, Norepinephrine, Depression, and Type 2 Diabetes. *Biomedicines*, *12*(6), 1187. 10.3390/biomedicines12061187

Porter, G. A., & O’Connor, J. C. (2022). Brain-derived neurotrophic factor and inflammation in depression: Pathogenic partners in crime? World Journal of Psychiatry, 12(1), 77–97. 10.5498/wjp.v12.i1.77

Poulter, J. A., Al-Araimi, M., Conte, I., van Genderen, M. M., Sheridan, E., Carr, I. M., Parry, D. A., Shires, M., Carrella, S., Bradbury, J., Khan, K., Lakeman, P., Sergouniotis, P. I., Webster, A. R., Moore, A. T., Pal, B., Mohamed, M. D., Venkataramana, A., Ramprasad, V., … Toomes, C. (2013). Recessive mutations in SLC38A8 cause foveal hypoplasia and optic nerve misrouting without albinism. American Journal of Human Genetics, 93(6), 1143–1150. 10.1016/j.ajhg.2013.11.002

Purcell, S., Neale, B., Todd-Brown, K., Thomas, L., Ferreira, M. A. R., Bender, D., Maller, J., Sklar, P., de Bakker, P. I. W., Daly, M. J., & Sham, P. C. (2007). PLINK: A Tool Set for Whole-Genome Association and Population-Based Linkage Analyses. The American Journal of Human Genetics, 81(3), 559–575. 10.1086/519795

Rahmani, M., Wong, B. W., Ang, L., Cheung, C. C., Carthy, J. M., Walinski, H., & McManus, B. M. (2006). Versican: Signaling to transcriptional control pathways. Canadian Journal of Physiology and Pharmacology, 84(1), 77–92. 10.1139/y05-154

Ransohoff, R. M., & Engelhardt, B. (2012). The anatomical and cellular basis of immune surveillance in the central nervous system. Nature Reviews Immunology, 12(9), 623–635. 10.1038/nri3265

Schroer, T. A. (2004). Dynactin. Annual Review of Cell and Developmental Biology, 20, 759–779. 10.1146/annurev.cellbio.20.012103.094623

Shaul, Y. D., & Seger, R. (2007). The MEK/ERK cascade: From signaling specificity to diverse functions. Biochimica Et Biophysica Acta, 1773(8), 1213–1226. 10.1016/j.bbamcr.2006.10.005

Shirayama, Y., Chen, A. C.-H., Nakagawa, S., Russell, D. S., & Duman, R. S. (2002). Brain-Derived Neurotrophic Factor Produces Antidepressant Effects in Behavioral Models of Depression. The Journal of Neuroscience, 22(8), 3251–3261. 10.1523/JNEUROSCI.22-08-03251.2002

Sprooten, E., Franke, B., & Greven, C. U. (2022). The P-factor and its genomic and neural equivalents: An integrated perspective. Molecular Psychiatry, 27(1), 38–48. 10.1038/s41380-021-01031-2

Takahashi, T., Fournier, A., Nakamura, F., Wang, L. H., Murakami, Y., Kalb, R. G., Fujisawa, H., & Strittmatter, S. M. (1999). Plexin-neuropilin-1 complexes form functional semaphorin-3A receptors. Cell, 99(1), 59–69. 10.1016/s0092-8674(00)80062-8

Thomas, G. M., & Huganir, R. L. (2004). MAPK cascade signalling and synaptic plasticity. Nature Reviews. Neuroscience, 5(3), 173–183. 10.1038/nrn1346

Tokuyama, W., Okuno, H., Hashimoto, T., Xin Li, Y., & Miyashita, Y. (2000). BDNF upregulation during declarative memory formation in monkey inferior temporal cortex. Nature Neuroscience, 3(11), 1134–1142. 10.1038/80655

Toyo-oka, K., Mori, D., Yano, Y., Shiota, M., Iwao, H., Goto, H., Inagaki, M., Hiraiwa, N., Muramatsu, M., Wynshaw-Boris, A., Yoshiki, A., & Hirotsune, S. (2008). Protein phosphatase 4 catalytic subunit regulates Cdk1 activity and microtubule organization via NDEL1 dephosphorylation. The Journal of Cell Biology, 180(6), 1133–1147. 10.1083/jcb.200705148

Vadhvani, M., Schwedhelm-Domeyer, N., Mukherjee, C., & Stegmüller, J. (2013). The centrosomal E3 ubiquitin ligase FBXO31-SCF regulates neuronal morphogenesis and migration. PloS One, 8(2), e57530. 10.1371/journal.pone.0057530

Ward, R. J., Zucca, F. A., Duyn, J. H., Crichton, R. R., & Zecca, L. (2014). The role of iron in brain ageing and neurodegenerative disorders. The Lancet. Neurology, 13(10), 1045–1060. 10.1016/S1474-4422(14)70117-6

Watanabe, K., Taskesen, E., Van Bochoven, A., & Posthuma, D. (2017). Functional mapping and annotation of genetic associations with FUMA. Nature Communications, 8(1), 1826. 10.1038/s41467-017-01261-5

Wendt, F. R., Pathak, G. A., Tylee, D. S., Goswami, A., & Polimanti, R. (2020). Heterogeneity and Polygenicity in Psychiatric Disorders: A Genome-Wide Perspective. Chronic Stress, 4, 247054702092484. 10.1177/2470547020924844

Xu, D., & Lu, W. (2020). Defensins: A Double-Edged Sword in Host Immunity. Frontiers in Immunology, 11, 764. 10.3389/fimmu.2020.00764

